# The efficiency in the ordinary hospital bed management in Italy: an in-depth analysis of intensive care unit in the areas affected by COVID-19 before the outbreak

**DOI:** 10.1101/2020.04.06.20055848

**Authors:** Fabrizio Pecoraro, Fabrizio Clemente, Daniela Luzi

## Abstract

In the first months of 2020 an increasing number of individuals worldwide are infected by the coronavirus disease 2019 (COVID-19). A particularly severe diffusion of the virus has affected Italy and in particular its northern regions. This is resulting in a high demand of hospitalization with a particular attention on the intensive care units (ICUs). Hospitals are suffering the high degree of patients to be treated for respiratory diseases and the majority of the structures located in the north of Italy are or are going to be saturated. This has led the actual and past national and regional governments to be heavily criticized for reducing in the past years the number of beds, in particular those located in the ICUs across the country. Aim of this study is to analyse the availability of hospital beds across the country as well as to determine their management in terms of complexity and performance of cases treated at regional level. The results of this study underlines that, despite the reduction of beds for the majority of the hospital wards, ICUs availabilities did not change between 2010 and 2017. Moreover, this study confirms that the majority of the Italian regions efficiently manage these structural facilities allowing hospitals to treat patients without the risk of having an overabundance of patients and a scarcity of beds. In fact, this analysis shows that, in normal situations, the management of hospital and intensive care beds has no critical levels.

## Introduction

The number of individuals infected with severe acute respiratory syndrome coronavirus 2 (SARS-CoV-2), the virus causing coronavirus disease 2019 (COVID-19), is dramatically increasing worldwide [1]. The first person-to-person transmission in Italy was reported on Feb 21st, 2020, and led to an infection chain that represents the largest COVID-19 outbreak outside Asia to date. At the moment, Italy is the second most affected country in the World and the first in Europe, with more than 62000 confirmed cases according to the Italian Department of Civil Protection as of 26th of March 2020. What it is not clearly identified so far is the reasons for the strong involvement of Italy within the European countries [2] and even more why the spread of this epidemic has not followed a uniform pattern on the territory: the diffusion of the disease, at the moment, is focused on the northern regions of the country. This was particularly evident considering that the virus took some days to reach the central and the southern regions of the land. Currently, the 87% of the confirmed actual cases are inhabitants of the northern regions that counts the 55% of the total Italian population. Among them, where the first cases were identified at the end February, is still the region with the majority of individuals infected by the COVID-19 virus. Similar results can be noted considering the hospitalization in general (88% in the northern regions and 42% in) and in the intensive care unit in particular (85% in the northern regions and 35% in). This data are provided and daily updated by the Italian Civil Protection Department [3]. These numbers are indicative of the spread of the infection and underline that one the main challenges to be faced by the National and the Regional Health Systems is the management of hospital resources across the country in terms of professionals, medical devices and hospital beds, with a particular attention on the intensive care unit (ICU) [4].

Given this high demand of hospitalization and critical care, Italian hospitals and in particular those located in the northern regions have been overloaded and the majority of them are struggling to cope with patients infected by the COVID-19 in addition to those who are hospitalized from other diseases. This has led the actual and past national and regional governments to be heavily criticized for reducing in the past years the number of beds, in particular those located in the ICUs across the country. Different newspapers have highlighted that a reduction in the number of beds have affected many hospitals over the territory. This resource reduction is mainly related to the progressive cutting of the national and local budget in the last decade that led the regional health services to close a significant number of local small dimension hospitals that generally have higher costs [5]. This lack of beds is confirmed by a recent study of the OECD that reported that our country counts 2.6 hospital beds per 1000 inhabitants [6], ranking Italy at the 19^th^ place over 23 countries with Germany having more than 6 beds per 1000 inhabitants. However, this data does not consider the number of ICUs. The most recent study reporting the number of beds specifically for the ICU was published by Rhodes et al in 2012 [7]. Also in this case the number of beds located in Italy are below the European average with 12.9 beds per 100000 inhabitants, while Germany is equipped with 29.2 beds. The analysis of the Italian National Healthcare Service should take into account that health services are organized and delivered under the responsibility of local authorities structured at a regional level, while the Italian Government has a weak strategic leadership.

Starting from these premises, the aim of this study is to analyse the Italian regional hospital systems in order to assess the efficiency and the performance of the hospital bed management in the years that precede the COVID-19 outbreak. Specific attention is given to the management of the intensive care unit departments. In this perspective, there are two methodologies adopted to assess the efficiency of a health structure in the management of clinical hospitalized cases: the first one is the hospital bed management [8-10] that provides an overall description of the use of beds by health structures. The second methodology evaluates the performance of a hospital considering the complexity of the cases treated by the structure [11-12]. Both methodologies investigate hospital performances providing a helpful snapshot for healthcare managers for the evaluation of healthcare systems [13].

The paper is structured as follows: after the materials and methods paragraph, a picture of the virus diffusion and workload of the hospital infrastructure is reported to provide an overview of the extraordinary event faced by the health and social care professionals in Italy. After that, a comparison of data captured in the years 2010 and 2017 is performed to verify the differences across years in terms of hospital beds available in each region. Finally, the results of the hospital bed management analysis are reported to capture the efficiency as well as the complexity and performance of patient hospitalizations.

## Materials and methods

### Data

This paper is focused on two main data sources. The first one provides daily data on the diffusion and hospital infrastructure consumption during the COVID-19 outbreak. They are collected by the Italian Civil Protection Department and collected on several web sites [3]. The second information flow concerns data on hospital bed management. Administrative and clinical data produced during the hospitalization process are collected in Discharge Report Forms and sent by the hospital information systems of a single health provider and centrally collected by the Ministry of Health. They describe each service provided to a patient as well as facilities available and staff employed in the relevant structure. This information is aggregated and published in the web site of the Ministry of Health [14]. The data analysed in this study refer to the year 2017, the latest data published by the Ministry of Health. Moreover, data captured for the year 2010 are analysed to examine differences between 2010 and 2017 in terms of resources available across the territory. This study considers both the public and private institutes, excluding the private nursing homes. The datasets gathered from the Ministry of Health website [14] and the relevant analyses performed during the current study are available in the Zenodo repository [15].

### Methodologies

The overall description of the hospital bed management is assessed using the following indicators computed on the basis of the number of beds, the patient discharged over a specific period of time (i.e. a year) and total number of in-patient days (i.e. the overall number of days that all the patients are hospitalized) [16].

- Beds Occupancy Rate (BOR): percentage of inpatient beds occupied over a specific period;
- Average Length Of Stay (AvLOS): average number of days that an inpatients remained in the hospital;
- Turnover Interval (TOI): number of days that an available bed remains empty between the discharge of a patient and the admission of a next one;
- Beds Turn Over (BTO): average number of patients “passing through” each bed during a specific period.

The authors also adopted reference thresholds for TOI (1<TOI<3) and BOR (BOR>75%). These values have been used to classify each region in the following areas: 1) the red one that identifies regions where both TOI and BOR are outside the reference threshold; 2) the yellow area that reports regions where either TOI or BOR are outside the threshold and 3) the green area that identifies regions where both indicators are within the reference thresholds.

The complexity and the performance of each clinical department in the management of clinical hospitalization cases are described, respectively, by the Case Mix (CMI) and the Performance Index (PI). In particular, the former indicates the degree of complexity in each observed unit with respect to a benchmark level (i.e. the national average), while the PI compares the performance of the observed unit considering the inpatient length of stays compared to the benchmark level.

These indicators are computed according to the following formulas, taking into account the different clinical specialties addressed by the relevant structure.

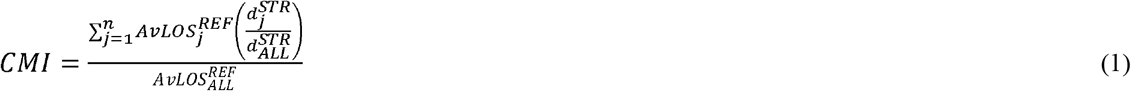

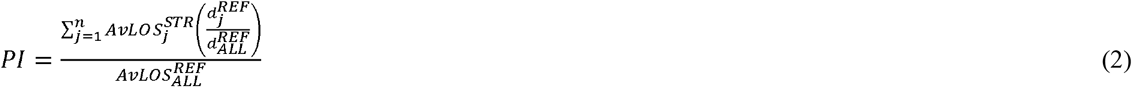

Where the variables indicate:

- *n*: number of specialties assessed by the structure;
- 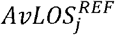: Italian AvLOS of the specialty j;
- 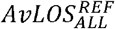: Italian AvLOS of all specialties;
- 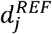: number of patient discharges in Italy of the specialty j
- 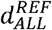: total number of patient discharges in Italy;
- 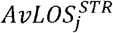: AvLOS of the specialty j in the relevant structure;
- 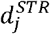: number of patients discharged in the relevant structure for the specialty j;
- 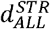: total number of patients discharges in the relevant structure.

A CMI (Equation 1) lower than 1 identifies a structure that manages low-level of complexity clinical cases, whereas a PI (Equation 2) lower than 1 identifies a structure with a high-level of efficacy. Considering the graphical representation, these indicators are analysed adopting a four-quadrant graph where the CMI is reported in the abscissa and the PI is reported in the ordinate compared to the national benchmark.

## Results

### Impact of the COVID-19 outbreak in the hospital bed management in Italy

Since the beginning of the infection through the identification of the first cases in the north of Italy, the number of patients infected closely follows an exponential trend, with a slightly change during the last week turning from an exponential to a linear trend. This tendency is appreciable not only considering the total number cases, but also the number of deaths, healed as well as the number of patients hospitalized in general and in the intensive care units dislocated all over the country. Trends describing the hospitalizations are reported in Figure 1 considering both the country as a whole and the region in particular, that still represents the region with the highest number of cases in Italy. These trends highlight the tremendous amount of work carried out by the healthcare professionals to host the exceptional number of patients to be hospitalized in the health structures.

**Figure 1.**
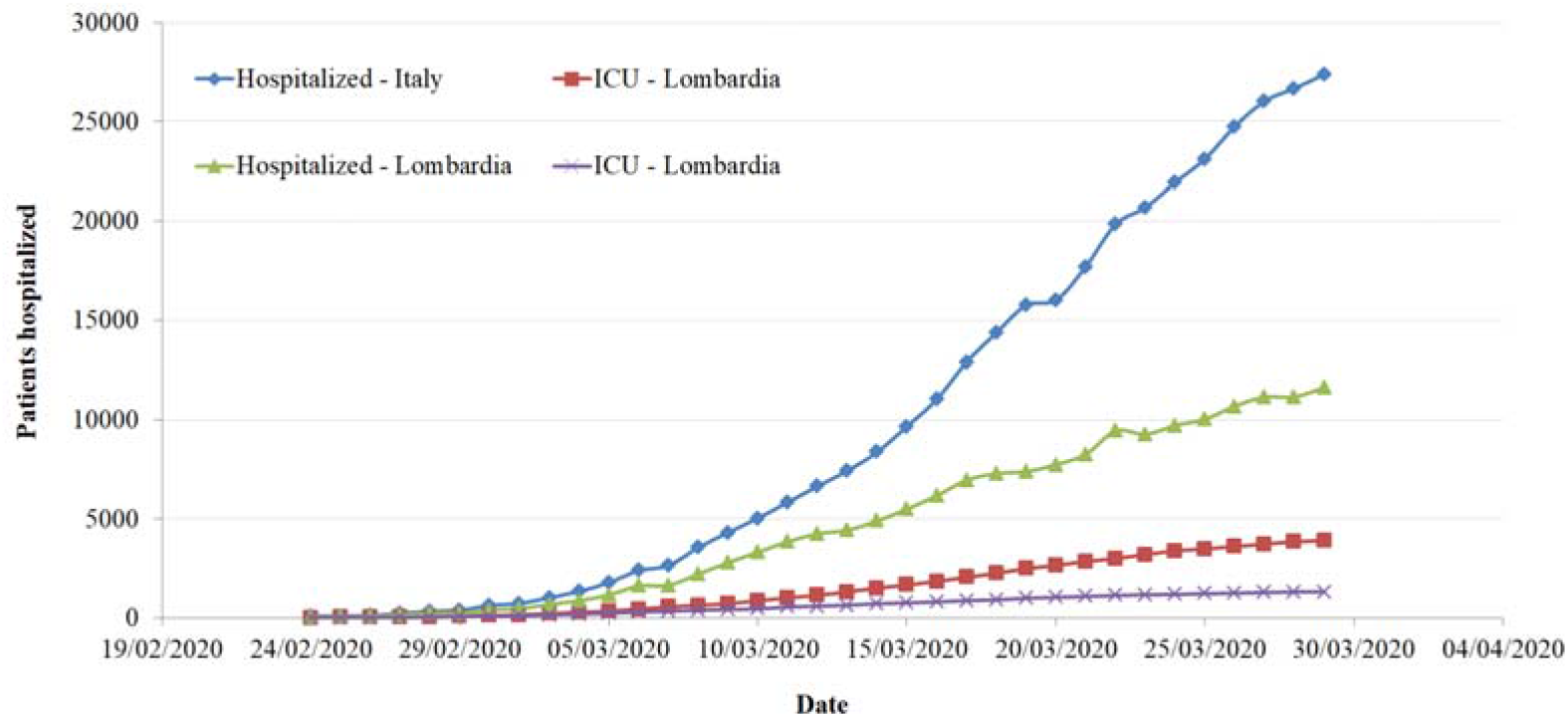
Number of patients hospitalized in the Italian and in the region structures in total as well as in the Intensive Care Units [3].

Although all hospital wards are struggling with an exceptional workload due to the COVID-19 outbreak, the intensive care units are particularly stressed given that the majority of the hospitals in the northern part of the country are saturating the hospital beds generally located in this department. Figure 2 summarized the actual occupancy rate of the ICU beds to date (29^th^ of March 2020). It is important to note that both the number of beds and the number of inpatients is purely indicative: the number of beds refer to the availability of each region in 2019 and does not take into account the epidemic, the number of patients hospitalized refers exclusively to patients affected by COVID-19 and does not take into account patients with other pathologies. As highlighted by the colour, different regions have had already saturated the hospital beds ordinarily available in their intensive care units (Lombardy, Piemonte, Val d’Aosta, Trento, Bolzano), with other regions that are worryingly approaching this threshold (Marche, Emilia Romagna, Toscana, Liguria). To be noted that, as highlighted in the map, all of them are located in the northern part of Italy.

**Figure 2.**
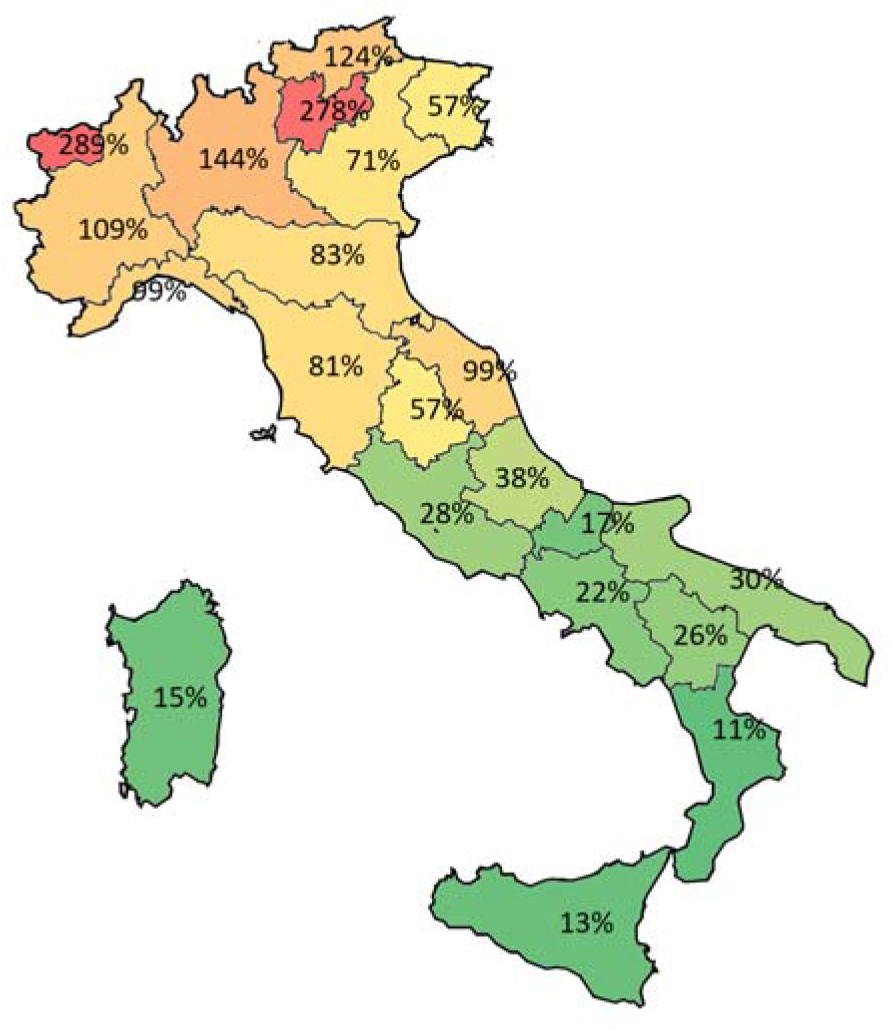
Percentage of beds occupied in the relevant regions considering the number of routinely available hospital beds and the number of patient hospitalized affected by COVID-19. Colours span from green to red depending on the percentage of beds occupied in the intensive care units.

### Analysis of the Italian structural components

In this paragraph data collected in the years 2010 and 2017 are compared to capture difference in terms of beds availability, number of hospitalizations and total number of inpatient days. A highlighted in Table 1, an important reduction of the hospital beds is evident all over the Italian regions. This reduction on the same time matches with a reduction of both number of patients hospitalized and number of days spent in the hospital. Differences across the country can be highlighted with the highest reductions in the southern regions. Considering the northern region heavily affected by the COVID-19, the reduction ranges from −5% in Bolzano and Val d’Aosta to −23% in Trento, while Lombardy cuts around the 7% of hospital beds.

**Table 1.**
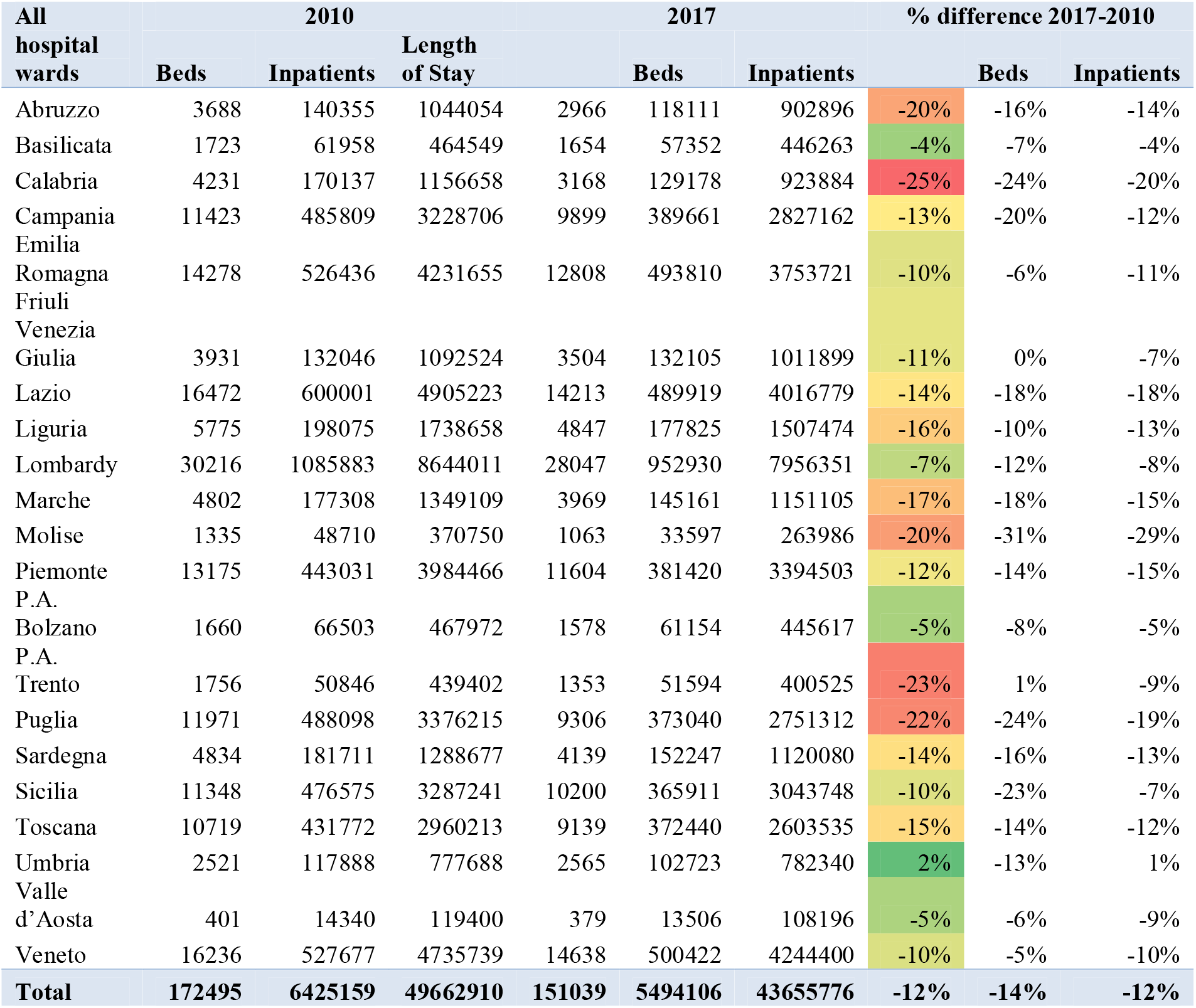
Number of hospital beds, hospitalization and length of stay in the years 2010 and 2017 highlighting the differences in each Italia region.

A different picture is shown focusing on the intensive care unit. Table 2 highlights that the availability of the number of beds in this department did not change across years with an increasing number of beds around 6% all over the territory. Of course, also in this analysis differences across regions can be detected. However, only five regions have reduced the number of beds in this department with the highest cut found in Lazio, Liguria and Piemonte (5%).

**Table 2.** Number of hospital beds, hospitalization and length of stay in the years 2010 and 2017 in the intensive care units highlighting the differences in each Italia region.

| Intensive care unit hospital ward | 2010 |  |  | 2017 |  |  | % difference 2017-2010 |  |  |
| --- | --- | --- | --- | --- | --- | --- | --- | --- | --- |
|  | Beds | Inpatients | Length of Stay | Beds | Inpatients | Length of Stay | Beds | Inpatients | Length of Stay |
| Abruzzo | 91 | 1105 | 18566 | 92 | 1198 | 17723 | 1% | 8% | -5% |
| Basilicata | 41 | 698 | 10576 | 49 | 745 | 9782 | 20% | 7% | -8% |
| Calabria | 107 | 1666 | 24931 | 124 | 2741 | 29237 | 16% | 65% | 17% |
| Campania | 363 | 7239 | 94649 | 427 | 6502 | 91616 | 18% | -10% | -3% |
| Emilia Romagna | 372 | 3710 | 53817 | 360 | 4201 | 51943 | -3% | 13% | -3% |
| Friuli Venezia Giulia | 103 | 1743 | 14424 | 119 | 1936 | 16132 | 16% | 11% | 12% |
| Lazio | 510 | 4770 | 98347 | 486 | 4699 | 91136 | -5% | -1% | -7% |
| Liguria | 173 | 1843 | 28332 | 164 | 1491 | 25157 | -5% | -19% | -11% |
| Lombardy | 663 | 6775 | 92057 | 738 | 7641 | 91661 | 11% | 13% | 0% |
| Marche | 113 | 1608 | 18885 | 127 | 1538 | 19826 | 12% | -4% | 5% |
| Molise | 29 | 413 | 6930 | 35 | 496 | 6980 | 21% | 20% | 1% |
| Piemonte | 316 | 3428 | 50239 | 299 | 3468 | 44595 | -5% | 1% | -11% |
| P.A. Bolzano | 36 | 616 | 7205 | 37 | 612 | 4592 | 3% | -1% | -36% |
| P.A. Trento | 20 | 272 | 2673 | 31 | 369 | 3628 | 55% | 36% | 36% |
| Puglia | 197 | 3891 | 53895 | 231 | 3884 | 56404 | 17% | 0% | 5% |
| Sardegna | 107 | 1614 | 25445 | 120 | 1695 | 22436 | 12% | 5% | -12% |
| Sicilia | 336 | 5240 | 73888 | 323 | 5118 | 76188 | -4% | -2% | 3% |
| Toscana | 322 | 3415 | 49959 | 373 | 3911 | 56092 | 16% | 15% | 12% |
| Umbria | 61 | 683 | 8615 | 69 | 730 | 8914 | 13% | 7% | 3% |
| Valle d'Aosta | 8 | 118 | 911 | 10 | 118 | 1273 | 25% | 0% | 40% |
| Veneto | 448 | 4851 | 68142 | 468 | 4777 | 64813 | 4% | -2% | -5% |
| **Total** | **4416** | **55698** | **802486** | **4682** | **57870** | **790128** | **6%** | **4%** | **-2%** |

A focus on the reduction of the hospital beds in the different wards is reported in Table 3 highlighting the disciplines that cover the 90% of the hospitalization complexity in terms of the average length of stay. As reported in the Table only oncology and intensive care unit wards have had an increase in the number of beds in Italy, while in other clinical specializations the reduction was even more than 20%, among which surgery, paediatrics and otolaryngology. This indicates that, among the wards that manage complex hospital cases, the availability of hospital beds was not affected by the funding cuts to public health. However, these crude differences may take into account that, in the last years, many European countries have adopted policies to strongly shift the organization and provision of health and social services from formal institutional facilities (e.g. hospitals) to home care [17]. Moreover, recently the provision of different scheduled procedures (e.g. diagnostic test, clinical examinations, treatments) are mainly provided in a day hospital, reducing the number of beds needed to treat the patients.

**Table 3.** Number of beds in the years 2010 and 2017 highlighting the differences in each hospital ward

| **Beds available in each hospital ward** | **2010** | **2017** | **% Difference 2010-2017** | **ICM reference value** |
| --- | --- | --- | --- | --- |
| General medicine | 30118 | 26053 | -13% | 0,20 |
| General surgery | 19854 | 15889 | -20% | 0,11 |
| Orthopedics and traumatology | 14314 | 12063 | -16% | 0,08 |
| Obstetrics and gynecology | 13000 | 10934 | -16% | 0,07 |
| Function recovery and rehabilitation | 10049 | 9708 | -3% | 0,06 |
| Cardiology | 6759 | 6687 | -1% | 0,05 |
| Long-term patients | 5135 | 4067 | -21% | 0,03 |
| Neurology | 5222 | 4944 | -5% | 0,03 |
| Pediatrics | 5746 | 4578 | -20% | 0,03 |
| Geriatrics | 4061 | 3550 | -13% | 0,03 |
| Urology | 5139 | 4501 | -12% | 0,03 |
| Psychiatry | 4172 | 4032 | -3% | 0,03 |
| Pneumology | 3777 | 3186 | -16% | 0,02 |
| Oncology | 2703 | 2759 | 2% | 0,02 |
| Infectious and tropical diseases | 3154 | 2816 | -11% | 0,02 |
| Neurosurgery | 2733 | 2454 | -10% | 0,02 |
| Intensive care | 4416 | 4682 | 6% | 0,02 |
| Otolaryngology | 3408 | 2549 | -25% | 0,02 |
| Neonatology | 1944 | 1913 | -2% | 0,01 |
| Nefrologia | 2049 | 1913 | -7% | 0,01 |
| Neuro-rehabilitation | 1981 | 1855 | -6% | 0,01 |
| Gastroenterology | 1716 | 1616 | -6% | 0,01 |

Other differences that can be detected analysing the availability of hospital beds refers to their distribution in the 21 regions and autonomous provinces. Figure 3 reports the number of beds per 100.000 inhabitants both considering all disciplines (A) and focusing on the intensive care unit departments (B). The proportions vary across the country spanning from Molise that counts the highest values for both categories to Calabria that displays the lowest proportions. In this overall picture, while, generally, the south of Italy suffers the lack of hospital beds, lays in the middle of the rank with 279 beds in total and 8 intensive care unit beds per 100000 inhabitants.

**Figure 3.**
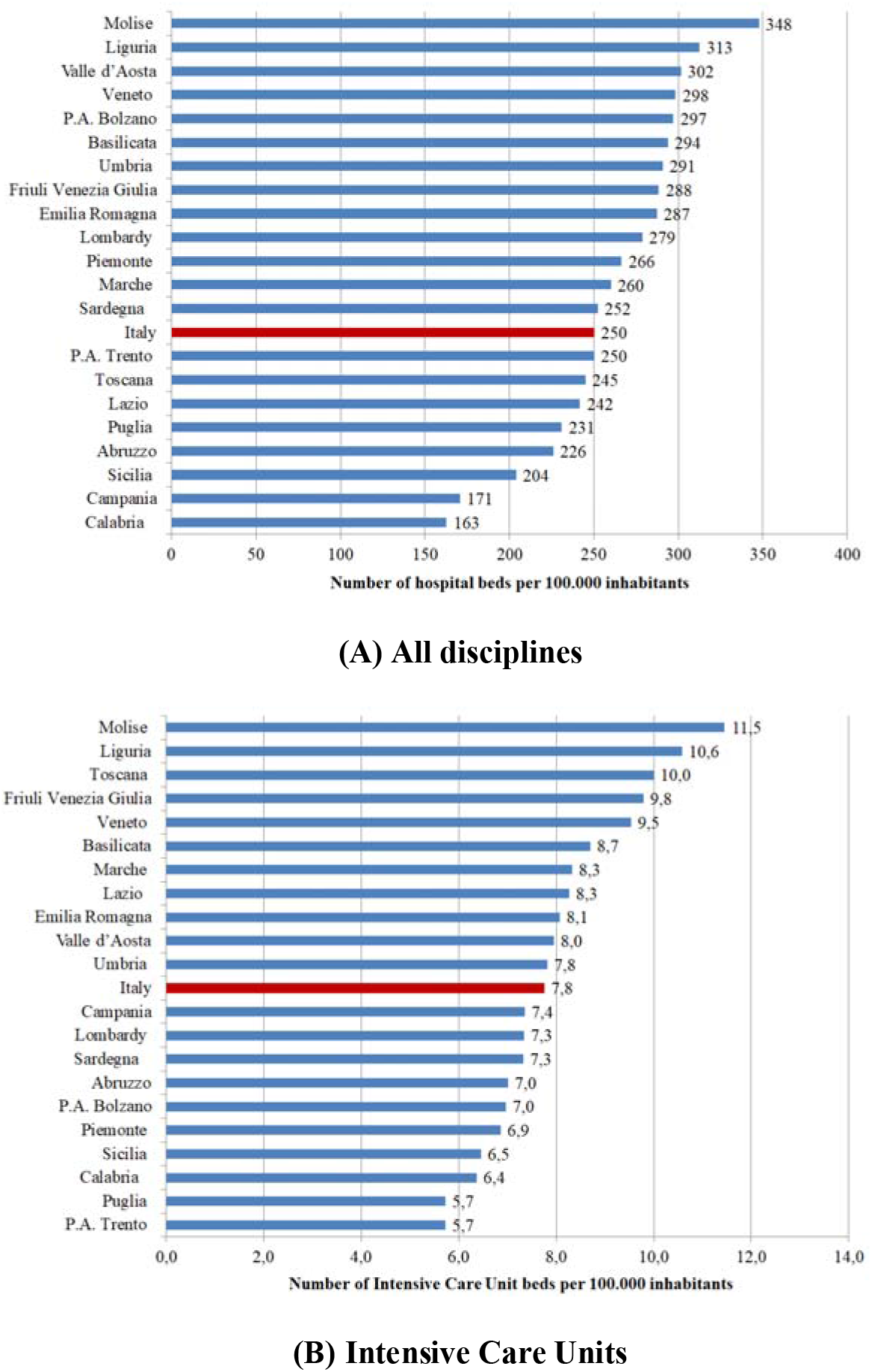
Number of total and intensive care unit beds per 100000 inhabitants in the different Italian regions.

### Hospital bed management

Table 4 reports the results obtained for each region highlighting the classification in the areas of hospital bed management as well as case-mix and performance indicators. Green cells highlight region where the indicators are above the relevant threshold. Note that while BOR and TOI thresholds are captured in the literature, those for the AvLOS and BTO are computed on the basis of the national average value. Considering the hospital bed management all regions fall within the threshold values both for the BOR and the TOI, with the exception of Molise, Basilicata and Sardegna. Instead, regional differences can be captured considering the AvLOS which highlights which are the regions that tend to extend the patient’s hospital stays comparing to a national benchmarking value (i.e. Lombardy and Liguria) and the BTO that provides an overview of the regions that have a low number of patients as a percentage of available beds (i.e. Lombardy and Molise). The difference in the AvLOS across the country can determine the level of performance of each region also on the basis of the complexity of cases. On the basis of these analyses, regions have been classified in the following four macro-clusters, that mainly represent the classification of the complex and performance indicators as reported in Figure 4:

**Table 4.** Cross-regional comparison of the results of bed management as well as complex and performance analysis

| All hospital wards | TOI | BOR | AvLOS | BTO | CMI | PI |
| --- | --- | --- | --- | --- | --- | --- |
| Abruzzo | 1,52 | 83% | 7,64 | 39,82 | 1,02 | 0,97 |
| Basilicata | 2,75 | 74% | 7,78 | 34,67 | 1,03 | 0,97 |
| Calabria | 1,80 | 80% | 7,15 | 40,78 | 0,95 | 0,98 |
| Campania | 2,02 | 78% | 7,26 | 39,36 | 0,94 | 0,99 |
| Emilia Romagna | 1,87 | 80% | 7,60 | 38,55 | 1,02 | 0,96 |
| Friuli Venezia Giulia | 2,02 | 79% | 7,66 | 37,70 | 1,01 | 1,00 |
| Lazio | 2,39 | 77% | 8,20 | 34,47 | 0,96 | 1,08 |
| Liguria | 1,47 | 85% | 8,48 | 36,69 | 1,08 | 1,01 |
| Lombardy | 2,39 | 78% | 8,35 | 33,98 | 1,03 | 1,04 |
| Marche | 2,05 | 79% | 7,93 | 36,57 | 0,96 | 1,06 |
| Molise | 3,69 | 68% | 7,86 | 31,61 | 1,03 | 1,02 |
| Piemonte | 2,20 | 80% | 8,90 | 32,87 | 1,03 | 1,10 |
| P.A. Bolzano | 2,13 | 77% | 7,29 | 38,75 | 0,99 | 0,93 |
| P.A. Trento | 1,81 | 81% | 7,76 | 38,13 | 1,01 | 0,98 |
| Puglia | 1,73 | 81% | 7,38 | 40,09 | 0,95 | 0,99 |
| Sardegna | 2,57 | 74% | 7,36 | 36,78 | 0,97 | 1,05 |
| Sicilia | 1,86 | 82% | 8,32 | 35,87 | 0,97 | 1,08 |
| Toscana | 1,97 | 78% | 6,99 | 40,75 | 1,00 | 0,91 |
| Umbria | 1,50 | 84% | 7,62 | 40,05 | 1,04 | 0,94 |
| Valle d' Aosta | 2,23 | 78% | 8,01 | 35,64 | 1,02 | 1,08 |
| Veneto | 2,20 | 79% | 8,48 | 34,19 | 1,03 | 1,04 |
| **Total** | **2,09** | **79%** | **7,95** | **36,38** |  |  |

**Figure 4.**
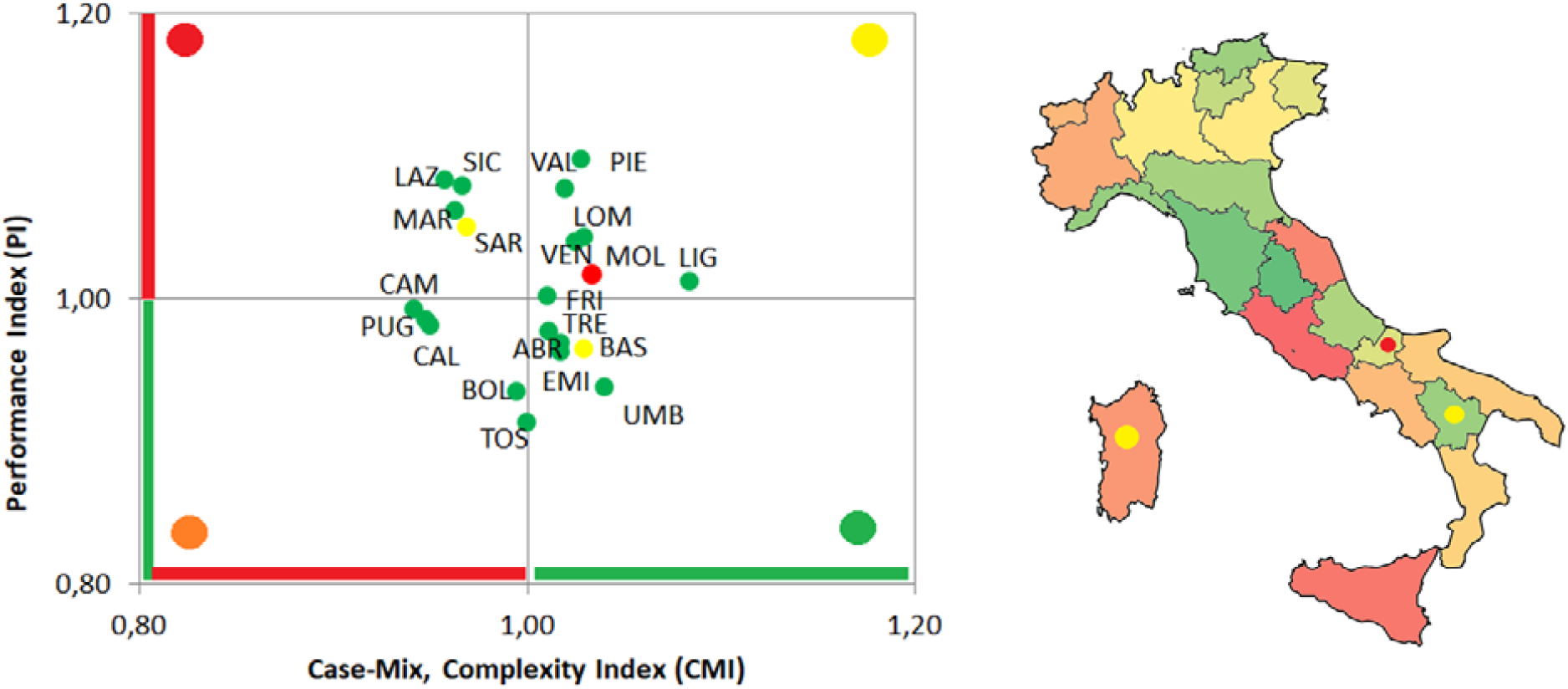
Overall classification of each Italian region considering both the hospital bed management indicators and the complexity and performance indicators. In both figures the colour of the marker captures the hospital bed management classification. In the Italian map, regions are coloured on the basis of the ratio between the CMI and PI.

1. Six regions (Abruzzo, Emilia Romagna, Trento, Toscana, Umbria e Basilicata) reported positive results in the management of hospital beds. Moreover, these regions report high performances in the treatment of complex cases, mainly due to short hospitalization with respect to the national average length of stay.
2. The second group of regions (Calabria, Campania, Bolzano e Puglia) has similar results compared to the above cluster, with hospitalizations efficiently managed with high performances. Differently from the above-mentioned group, these regions tend to manage cases that are less complex.
3. Six regions (Friuli Venezia Giulia, Piemonte, Valle D’Aosta, Veneto, Liguria e Lombardy) manage the hospital beds with a high turnover and beds that remain scarcely empty during the year. However, this is mainly due to a high length of stay that result in a low level of performance with respect to the national level, even if these regions mainly manage complex cases.
4. The last group is composed by four regions (Lazio, Marche, Sicilia e Sardegna). Even if they have positive values in the hospital bed turnover and turnover interval, the performance is lower than the national reference value also considering that these regions tend to manage less complex cases.

In this classification the Molise represents an interesting outlier given that it is the only region with both BRO and TOI outside the efficiency thresholds. These features contribute also to the performance of the regional hospitals even if this region tend to manage complex cases.

The analysis reported above is presented in the following, taking into account exclusively the intensive care unit beds and hospitalization flow. Table 5 reported the results of the hospital bed management, complexity and performance indicators. Also in this case, green cells identify efficient regions. In this analysis the thresholds for the four hospital bed management indicators are all defined on the basis of the national average values, given that no reference values are available in the literature. The macro-clusters detected on the basis of both analyses are summarized in the following (Figure 5):

**Table 5.** Cross-regional comparison of the results of bed management as well as complex and performance analysis of intensive care units.

| Intensive Care Unit | TOI | BOR | AvLOS | BTO | CMI | CPI |
| --- | --- | --- | --- | --- | --- | --- |
| Abruzzo | 13,24 | 53% | 14,79 | 13,02 | 0,97 | 1,14 |
| Basilicata | 10,88 | 55% | 13,13 | 15,20 | 1,25 | 1,02 |
| Calabria | 5,85 | 65% | 10,67 | 22,10 | 2,05 | 0,81 |
| Campania | 9,88 | 59% | 14,09 | 15,23 | 1,58 | 1,04 |
| Emilia Romagna | 18,91 | 40% | 12,36 | 11,67 | 0,81 | 0,91 |
| Friuli Venezia Giulia | 14,10 | 37% | 8,33 | 16,27 | 1,39 | 0,62 |
| Lazio | 18,36 | 51% | 19,39 | 9,67 | 0,91 | 1,42 |
| Liguria | 23,27 | 42% | 16,87 | 9,09 | 0,80 | 1,24 |
| Lombardy | 23,26 | 34% | 12,00 | 10,35 | 0,76 | 0,88 |
| Marche | 17,25 | 43% | 12,89 | 12,11 | 1,01 | 0,95 |
| Molise | 11,68 | 55% | 14,07 | 14,17 | 1,41 | 1,14 |
| Piemonte | 18,61 | 41% | 12,86 | 11,60 | 0,86 | 0,95 |
| P.A. Bolzano | 14,56 | 34% | 7,50 | 16,54 | 0,96 | 0,60 |
| P.A. Trento | 20,83 | 32% | 9,83 | 11,90 | 0,68 | 0,75 |
| Puglia | 7,19 | 67% | 14,52 | 16,81 | 0,98 | 1,08 |
| Sardegna | 12,60 | 51% | 13,24 | 14,13 | 1,05 | 0,98 |
| Sicilia | 8,15 | 65% | 14,89 | 15,85 | 1,33 | 1,09 |
| Toscana | 20,47 | 41% | 14,34 | 10,49 | 1,00 | 1,05 |
| Umbria | 22,29 | 35% | 12,21 | 10,58 | 0,67 | 0,91 |
| Valle d'Aosta | 20,14 | 35% | 10,79 | 11,80 | 0,85 | 0,87 |
| Veneto | 22,19 | 38% | 13,57 | 10,21 | 0,90 | 1,00 |
| **Total** | **15,88** | **46%** | **13,65** | **12,36** |  |  |

**Figure 5.**
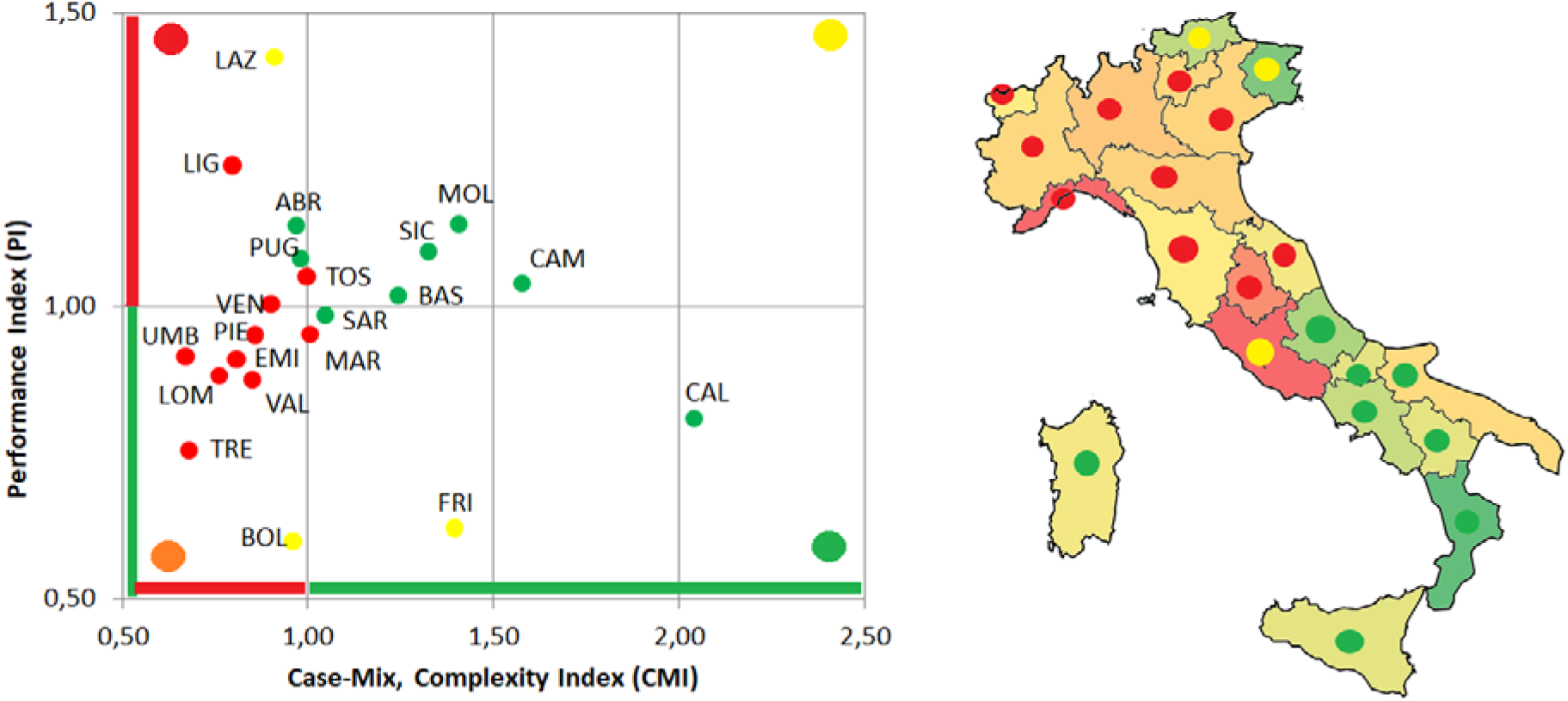
Overall classification of each Italian region considering both the hospital bed management indicators and the complexity and performance indicators of intensive care units. In both figures the colour of the marker captures the hospital bed management classification. In the Italian map, regions are coloured on the basis of the ratio between the CMI and PI.

1. In three regions (Calabria, Sardegna e Sicilia) all indicators report positive values with respect to the national average ones. They manage complex cases with high performance results. Moreover, the quick turnover and a high percentage of bed occupancy make these regions efficient also in the management of hospital beds.
2. A consistent number of regions (Basilicata, Campania, Molise, Sicilia, Abruzzo, Puglia e Lazio) belong to the second cluster. As reported for the previous group also these regions have a high bed turnover and the number of days between two hospitalization are relatively low. However, differently from the above mention group, these results are mainly due to the high number of days that patients spend in the intensive care unit. Also for this reason, even if these regions tend to cope with complex cases and pathologies, their performance is lower than the national one.
3. In three regions (Liguria, Toscana e Veneto) hospital beds are not efficiently managed with a low performance in the management of not so complex cases. This is mainly due to the low complexity of cases hospitalized with a length of stay higher than the national average value.
4. In the last group of regions (Emilia Romagna, Lombardy, Piemonte, Trento, Umbria, Val D’Aosta, Bolzano) the majority of the hospitalization cases are not efficiently managed with a high value of length of stay for not complex cases.

In this classification Marche represents an outlier in the management of hospital beds. Differently from the last reported cluster it manages complex cases with a relative high performance.

## Discussion and conclusions

COVID-19 is one of the most serious respiratory pandemics of the past 100 years, its rapid global spread is overwhelming hospitals and local communities in Italy. One of the main challenges to be faced by the Italian hospitals is to provide hospital beds for patients affected by this virus with a particular attention of the intensive care unit department. This is particularly evident considering that, at the moment, the majority of the northern regions in Italy are suffering with a low number of available beds and that others presumably saturate their capacity in the following days.

The lack of hospital beds has been attributed to a set of important cuts in the financing of public health over the last years that also result in a reduction of hospital beds both in general and in the intensive care units on the whole territory.

This paper provides an overview of the efficiency in the management of hospital beds across Italian regions also highlighting whether a substantial reduction of hospital beds has been carried out recently. The results of the overall analysis firstly show that even if an important reduction of hospital beds affects the majority of the hospital wards, an opposite trend has been detected for the intensive care unit. This is particularly evident considering the regions that are most effected by the virus: Lombardy +11%, Marche +12%, Piemonte −5%, Trento +55%, Bolzano +3%, Veneto +4%.

Hospital beds are generally efficiently managed all over the country with some exceptions in the south of Italy that show a slow turnover and/or a low bed occupancy rate. This positive result is particularly evident in the regions of northern Italy where cases are handled with rapid shifts, also without leaving beds empty during the year. Moreover, generally northern regions mainly deal with complex cases albeit with a performance below the national average. On the contrary, different results are displayed analysing the management of beds in the intensive care units, where the 75% occupancy rate threshold is not reached in all regions. In this sense, the northern regions exhibit a high performance, even if related to the management of less complex cases.

In conclusion this study highlights that in the recent years there has been no substation reduction of the hospital beds located in the intensive care units. Differently other wards suffered of the financial cuts done by national and regional governments to the public health. However, this trend may also take into account two important factors. On the one hand, the provision of health services is now shifting from formal institutional facilities (e.g. hospitals) to home care and, on the other hand, different scheduled procedures are mainly provided in a day hospital reducing the number of beds needed to treat the patients. The majority of the Italian regions and in particular those in the northern part of the country can rely on an appropriate number of beds that generally do not saturate in periods when the patient flow is not conditioned by catastrophic events like what we have been going through these weeks. This availability of hospital beds as well as the efficiency in their management confirmed in this study allow hospitals to treat patients without the risk of having an overabundance of patients and a scarcity of beds. In fact, the analysis reported in this paper showed that, in normal situations, the management of hospital and intensive care beds has no critical levels, while in pandemic or other catastrophic periods, the hospital management paradigms change [18] making it necessary also to change the relationship between hospital and territory as well as to determine the appropriate allocation of in-patient resources [19]. Considering that post-intensive care and intensive care continuously bed availability daily changes across the land and the European countries, it is not possible to apply this methodology in this pandemic period. However, an ex post analysis should be applied in the future to provide indications on the hospital-territory relationship in response to widespread social problems.

## Data Availability

The source datasets gathered from the Ministry of Health website and the relevant analyses performed during the current study are available in the Zenodo repository identified with the doi: 10.5281/zenodo.3737840

https://zenodo.org/record/3737840

## Notes

### Competing Interest Statement

The authors have declared no competing interest.

### Funding Statement

This research received no specific grant from any funding agency in the public, commercial, or not-for-profit sectors.

